# Serum neurofilament light chain predicts long-term prognosis in Guillain-Barré syndrome patients

**DOI:** 10.1101/2020.03.24.20042200

**Authors:** Lorena Martín-Aguilar, Pol Camps-Renom, Cinta Lleixà, Elba Pascual-Goñi, Jordi Diaz-Manera, Ricard Rojas-García, Noemí de Luna, Eduard Gallardo, Elena Cortés-Vicente, Laia Muñoz, Daniel Alcolea, Alberto Lleó, Carlos Casasnovas, Christian Homedes, Gerardo Gutiérrez-Gutiérrez, María Concepción Jimeno-Montero, José Berciano, Maria José Sedano Tous, Tania Garcia-Sobrino, Julio Pardo-Fernandez, Celedonio Márquez-Infante, Iñigo Rojas-Marcos, Ivonne Jericó Pascual, Eugenia Martínez-Hernández, Germán Morís de la Tassa, Cristina Domínguez-González, Isabel Illa, Luis Querol

## Abstract

**Objective:** To study baseline serum neurofilament light chain (sNfL) levels as a prognostic biomarker in Guillain-Barré syndrome (GBS).

**Methods:** We measured NfL using SiMoA in serum (98 samples) and CSF (24 samples) of GBS patients prospectively included in the International GBS Outcome Study (IGOS) in Spain and compared them with controls (HC). We performed multivariable regression to analyze the association between sNfL levels and functional outcome at one year.

**Results:** GBS patients had higher NfL levels than HC in serum (55.49pg/mL vs 9.13pg/mL, p<0,0001) and CSF (1308.5pg/mL vs 440.24pg/mL, p=0.034). Patients with preceding diarrhea had higher sNfL than patients with respiratory symptoms or no preceding infection (134.90pg/mL vs 47.86pg/mL vs 38.02pg/mL, p=0.016). sNfL levels correlated with GDS and R-ODS scales. Patients with pure motor variant and Miller-Fisher syndrome showed higher sNfL levels than patients with sensory-motor GBS (162.18pg/mL vs 95.50pg/mL vs 38.02pg/mL; p=0.025). AMAN patients had higher sNfL levels than other variants (190.55pg/mL vs 46.79pg/mL, p=0.013). sNfL returned to normal levels at one year. High baseline sNfL levels were associated with inability to run (OR=1.65, 95% CI 1.14-2.40, p=0.009) and lower R-ODS (β −2.60, 95% β −4.66-(−0.54), p=0.014) at one year. Cut-off points predicting clinically relevant outcomes at one year with high specificity were calculated: inability to walk independently (>319pg/mL), inability to run (>248pg/mL) and ability to run (<34pg/mL).

**Conclusion:** Baseline sNfL levels are increased in patients with GBS, they are associated with disease severity and axonal variants and they have an independent prognostic value in GBS patients.

## INTRODUCTION

Guillain-Barré syndrome (GBS) diagnosis relies on clinical and electrophysiological criteria^1^ and albumino-cytological dissociation in CSF, but patients differ considerably in their presentation, clinical course and prognosis. Previous prognostic models, based on clinical and epidemiological features, predict the ability to walk at six months^2,3^ or the probability to develop respiratory insufficiency^4^.

Neurofilament light chain (NfL) is becoming the most important axonal damage biomarker^5,6^ in Neurology after the introduction of ultrasensitive techniques such as the single-molecule array (SiMoA)^7,8^.

Older studies investigated the role of neurofilament heavy chain (NfH), showing increased CSF NfH levels in GBS patients^9,10^. CSF NfL levels were also described to associate with outcome in a small study^11^. A recent study showed increased serum and CSF NfL levels in acquired peripheral neuropathies, including five GBS patients^12^ and another study showed increased sNfL levels in various neurodegenerative diseases, including 19 GBS patients, but clinical correlations were not performed^13^. A very recent retrospective study showed that high sNfL were associated with poor short-term prognosis, but prospective or long-term prognosis data and the role of confounding variables were not studied^14^. The role of sNfL measured by SiMoA as a long-term prognosis biomarker in GBS has not been investigated so far.

Our study aims to study: (1) the levels of NfL in serum and CSF in GBS patients and its variants; (2) the kinetics of sNfL-at baseline and at one year; and (3) the relationship between baseline sNfL levels and prognosis at one year.

## MATERIALS AND METHODS

### Subjects, Standards Protocol Approvals and Patient Consents

We collected data, involving 98 patients enrolled in the Spanish cohort of the International Guillain-Barré Syndrome Outcome Study (IGOS) study^15^. The IGOS is a prospective, observational cohort study including all patients within the GBS diagnostic spectrum. Patients fulfilling diagnostic criteria for GBS or its variants were included within 2 weeks from onset; there were no exclusion criteria. Patients from the Spanish cohort were enrolled between February 2013 and January 2019. Additionally, 16 age-matched healthy controls for sera and 10 age-matched healthy controls for CSF were included. Serum and CSF samples were aliquoted and stored at −80°C until needed. The study was approved by the review boards of Erasmus University Medical Centre, Rotterdam, The Netherlands, the Ethics Committee of the Hospital de la Santa Creu i Sant Pau and the local institutional review boards of participating hospitals or universities. All patients gave written informed consent to participate in the study according to the Declaration of Helsinki.

### SiMoA NfL measurements

Measurement of serum and CSF NfL levels was performed in duplicates in all available samples from GBS patients and healthy controls. The analysis was performed using the reagents by investigators blinded to clinical data using SIMOA Nf-light® kit in SR-X immunoassay analyzer, Simoa™(Quanterix Corp, Boston, MA, USA), which runs ultrasensitive paramagnetic bead-based enzyme-linked immunosorbent assays. Samples were analyzed following the manufacturer’s instructions and standard procedures. To analyze the kinetics of serum NfL in GBS patients, we measured sNfL levels in those patients with available sample at the 52 weeks timepoint (n=33). All NfL values were within the linear ranges of the assays. The intra- and inter-assay coefficients of variation (ICV) at intermediate level (16.15pg/ml) were 7.1% and 6.3%, respectively.

### Data collection

Data were collected prospectively, regarding demography (age and gender), evolution time (days from clinical onset), infectious antecedent event and disability by the GBS disability score^16^ (GDS) at study entry. Results of routine CSF examination, nerve conduction studies (NCS) and treatment regimens were collected. We defined an elevated CSF protein level as higher than 0,45g/L^15,17^. Clinical variants were defined as sensorimotor, pure motor, pure sensory, Miller-Fisher syndrome (MFS), ataxic and pharyngeal-cervical-brachial variant^18^. Patients were classified as acute inflammatory demyelinating polyneuropathy (AIDP), acute motor axonal neuropathy (AMAN), acute motor-sensory axonal neuropathy (AMSAN), equivocal or normal based on initial NCS. We recorded GBS disability score (GDS) initially, at 4 weeks, at 26 weeks and at 52 weeks and its maximum score during follow-up. Rasch-built Overall Disability Scale (R- ODS, highest score 48, indicating no disability)^19^ was assessed at 4 weeks, 26 weeks and 52 weeks. Ability to run was extracted from R-ODS, and ability to walk independently was extracted from GDS.

### Statistical analysis

Descriptive statistics are showed as mean (+/- standard deviation) or median (interquartile range – IQR) in continuous variables and as frequencies (percentages) in categorical variables. sNfL levels were non-normally distributed as tested by the Shapiro-Wilk normality test. Thus, a logarithmic transformation of the variable was performed to approach the normal distribution (logNfL). We summarized NfL levels using geometric means (GeoMeans), GeoMean 95% confidence interval (CI) and coefficient of variation (CV). Comparisons between patients with GBS and healthy controls (HC) were performed by the Wilcoxon rank sum test. Kruskall-Wallis test was used to compare groups at baseline. Wilcoxon matched pairs signed rank test was used to compare baseline sNfL and sNfL at 52 weeks. We used Spearman’s coefficient to assess correlation between variables.

To investigate the association between sNfL and prognosis we performed two types of multivariable regression analyses. First, we conducted a multivariable logistic regression analysis to predict the ability to run at 1 year of follow-up. Secondly, we performed a multivariable linear regression analysis to investigate the association between logNfL and R-ODS scale at 1 year. In both analyses we performed a stepwise backward regression modeling to select variables independently associated with the outcome. The variables introduced in our initial multivariable models were selected based on known prognostic factors (age, GDS, diarrhea and AMAN^20,21^). To perform the multivariable analysis we excluded patients with MFS, because our aim was to predict GBS prognosis and MFS is considered a different disease, including different pathophysiology, clinical presentation (it does not present with weakness), treatment (often untreated) and outcome (considered self-limiting and benign). The final models were adjusted by potential confounders. A significant confounding effect was defined as an absolute change >10% in the regression coefficients when introducing the variable into the model.

Odds-ratios (OR) for the logistic regression analysis and beta coefficients (β) for the linear regression analysis were reported with 95% confidence intervals and p values.

Additionally, we evaluated the predictive capacity of NfL levels in the acute phase and at one year (residual disability) for 5 different clinically relevant endpoints, stablished prior to the analysis: (1) Ability to run at 1 year, (2) Inability to run at 1 year, (3) Inability to walk without assistance at 1 year, (4) Ventilation, and (5) Death. First, we performed a univariate logistic regression analysis for each endpoint using NfL levels as the exposure variable. Then, we performed a ROC analysis comparing the predictions against the endpoint and eventually, we selected the cut-off points of NfL levels that better predicted the endpoint aiming for the highest specificity. Finally, we tested the predictive capacity of each cut-point using multivariable logistic regression analyses adjusting for age and AMAN as possible confounders. OR with 95% CI, area under the curve (AUC), sensitivity, specificity, positive predictive value (PPV) and negative predictive value (NPV) for each cut-point were reported.

Statistical significance for all analyses was set at 0.05 (two-sided). The analysis was carried out in Graphpad Prism v8 and Stata v15® (Texas, USA).

### Data availability

Anonymized data not published within this article will be made available by request from any qualified investigator.

## RESULTS

### Baseline characteristics

We enrolled 98 participants from 11 Spanish centres participating in the IGOS study. GBS patients had an average of 57.4 years and were predominantly men (57%). 68.4% of patients presented an antecedent infectious event, mainly upper respiratory tract infections (43.9%) and diarrhea (24.5%). The median evolution time from onset of symptoms to inclusion was 4 days (IQR 3-6). Most patients had an initial GDS between 2 and 4 (93.9%), with a median of 3. CSF was examined in 90 (91.8%) patients within a median time of 3 days (IQR 2-6) from onset of neurological symptoms. Elevated CSF protein level was detected in 68.9% of the patients. 18.4% of patients presented with a pure motor GBS variant, 10.2% with MFS, 4.1% with pure sensory variant and 1 patient with ataxic variant; 66.3% presented with the typical sensory-motor variant. Regarding EMG classification, most patients had an AIDP (59.2%), followed by equivocal NCS (14.3%), AMAN (12.2%) and AMSAN (7.1%). Seven patients had normal NCS. Most patients were treated with IVIg (77.6%) or IVIg plus plasma exchange (10.2%). 5% of patients received a second course of IVIg and six patients did not receive treatment.

### Association of baseline sNfL with disease characteristics

GBS patients had significantly higher serum NfL (sNfL) levels (55.49pg/mL) than healthy controls (HC) (9.13pg/mL, p<0,0001, **table 1, figure 1A**). This difference was also observed in CSF (1308.5pg/mL vs 440.24pg/mL, p=0.034, **figure 1B**). We found a correlation between sNfL and CSF NfL (r=0.62, p<0.001, **supplementary figure. 1A**).

**Table 1.** Baseline serum and CSF NfL in patients with Guillain-Barré (GBS) and healthy controls (HC)

|  |  | GBS patients | HC | p |
| --- | --- | --- | --- | --- |
| Age | mean +/- SD | 57.4 +/- 16.6 | 51.7 +/- 19.4 | 0.173 |
| Gender | n. % male | 56 (57.1%) | 13 (50%) | 0.51 |
| Serum NfL (pg/mL) | n | 98 | 16 | <0.001 |
|  | GeoMean | 55.49 | 9.13 |  |
|  | Median | 40.00 | 8.22 |  |
|  | Max | 3573.03 | 22.82 |  |
|  | Min | 4.22 | 3.29 |  |
| CSF NfL (pg/mL) | n | 24 | 10 | 0.034 |
|  | GeoMean | 1308.50 | 440.24 |  |
|  | Median | 883.57 | 493.79 |  |
|  | Max | 32915.47 | 1469.97 |  |
|  | Min | 212.74 | 139.42 |  |

GBS = Guillain-Barré Syndrome; HC = Healthy controls; NfL = neurofilament light chain; CSF= cerebrospinal fluid. GeoMean = Geometric mean; Max = maximum; Min = minimum; SD= Standard Deviation.

**Figure 1.**
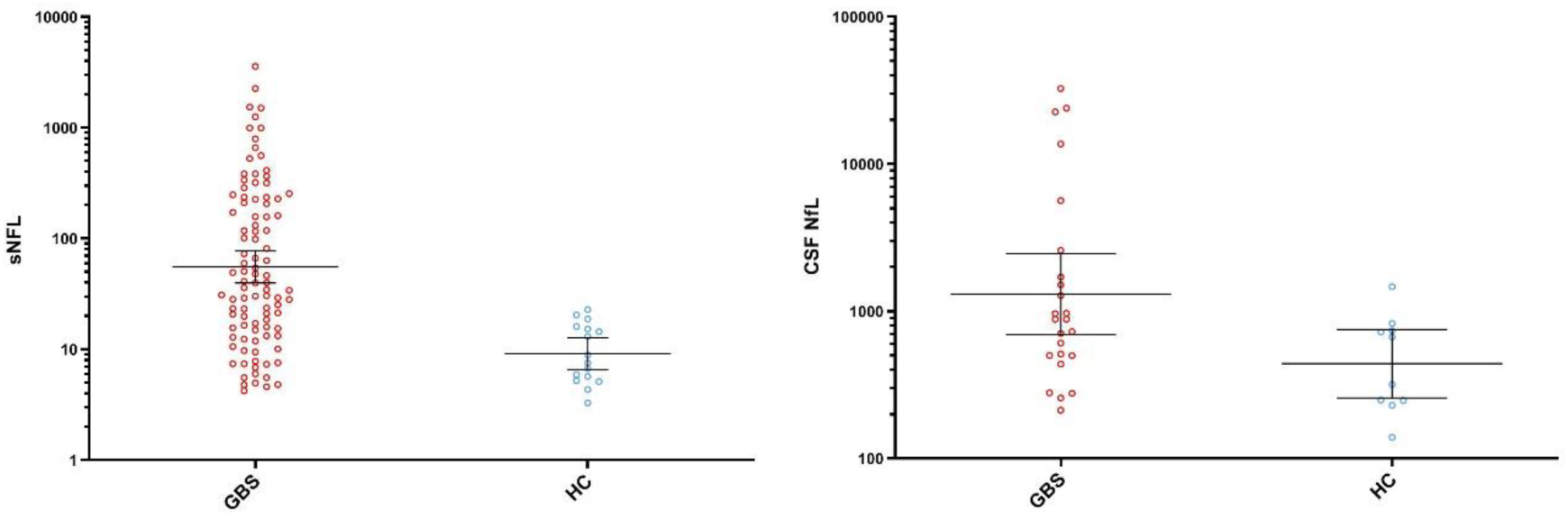
Baseline NfL in patients with Guillain-Barré (GBS) vs. healthy controls (HC) in serum (A) and in CSF (B). The line in the center represents the GeoMean value and the whiskers indicate the 95% confidence interval. GBS = Guillain-Barré Syndrome; HC = Healthy controls; sNfL = serum neurofilament light chain; CSF= cerebrospinal fluid. GeoMean = Geometric mean.

We did not find a correlation between sNfL levels and age in GBS patients (r=0.18, p=0.07), although it is clear in HC (r=0.65, p<0.001) (**supplementary figure 1B**). Patients with diarrhea had higher sNfL levels than those patients without any infectious antecedent or with respiratory symptoms (134.9pg/ml vs 38.02pg/mL vs 47.86pg/mL, respectively, p=0.016). CSF protein levels were not correlated with sNfL or CSF NfL. sNfL and CSF NfL levels were not correlated with time since symptom onset.

Regarding GBS variants, sNfL levels were higher in patients with pure motor variant and MFS comparing them with typical GBS (162.18pg/mL vs 95.5pg/mL vs 38.02pg/mL, respectively; p=0.025; **figure 2A**). When we stratified patients according to NCS, patients with AMAN had higher sNfL levels than other patients (199.53pg/mL vs 46.77pg/mL, p=0.006) and patients with equivocal NCS and AMSAN had higher sNfL levels than AIDP (**figure 2B**). The seven patients with normal NCS showed higher sNfL levels than AIDP patients (53.7pg/mL vs 35.48pg/mL, **table 2**). This result can be explained because five of these patients with normal NCS are MFS, who have higher sNfL than AIDP patients. When we stratified these patients according to clinical variants we found five MFS patients, one patient with pure sensory variant and one patient without clinical variant (89.13pg/mL vs 10.47pg/mL vs 23.44pg/mL, respectively). Patients who did not receive treatment (6.12%) had normal levels of sNfL. Further information about sNfL levels and disease characteristics is detailed in **table 2**.

**Table 2.** Relationship between sNfL levels and basal characteristics

| **BASAL CHARACTERISTICS (n=98)** |  | **sNfL (pg/ml; GeoMean.**<br>**GeoMean 95% CI. CV%)** | **p** |
| --- | --- | --- | --- |
| Age (mean +/- SD) | 57.35 +/-16.6 | - | 0.071 |
| Gender (n. %) |  |  |  |
| Male | 56 (57.1%) | 52.48 (31.62; 79.43). 41.5% | 0.625 |
| Female | 42 (42.9%) | 60.26 (39.81; 100). 41.8% |  |
| Infectious antecedent (n. %) |  |  |  |
| Diarrhea | 24 (24.5%) | 134.90 (63.10; 251.19). 36.6% | 0.016 |
| Respiratory infection | 43 (43.9%) | 38.02 (25.12; 63.10). 42.6% |  |
| None | 31 (31.6%) | 47.86 (28.35; 81.50). 39.3% |  |
| Evolution time (days. median; IQR) | 4 (3-6) | - | 0.413 |
| Initial GDS (median; IQR) | 3 (2-4) |  |  |
| 1 (n. %) | 4 (4.1%) | 6.17 (3.98; 10). 25.4% | 0.005 |
| 2 (n. %) | 28 (28.6%) | 35.48 (19.95; 63.10). 46.1% |  |
| 3 (n. %) | 19 (19.4%) | 64.56 (31.62; 125.89). 36.5% |  |
| 4 (n. %) | 45 (25.9%) | 77.62 (59.12; 125.89). 35.7% |  |
| 5 (n. %) | 2 (2%) | 375.83; 53.5% |  |
| GBS Variants (n. %) |  |  |  |
| No | 65 (66.3%) | 38.02 (25.12; 50.12); 37% | 0.009 |
| Pure motor | 18 (18.4%) | 162.18 (79.43; 398.11); 34.7% |  |
| Pure sensory | 4 (4.1%) | 26.30 (1.26; 501.19); 91.6% |  |
| MFS | 10 (10.2%) | 95.50 (31.62; 316.23); 42.3% |  |
| Ataxic | 1 (1%) | 338.84 |  |
| EMG Classification (n. %) |  |  |  |
| AIDP | 58 (59.2%) | 33.88 (25.12; 50.12). 37.7% | 0.008 |
| AMAN | 12 (12.2%) | 199.53 (79.43; 501.19); 28.5% |  |
| AMSAN | 7 (7.1%) | 107.15 (25.12; 501.19); 44% |  |
| Equivocal | 14 (14.3%) | 104.7 (39.81; 316.22); 43% |  |
| Normal | 7 (7.1%) | 53.70 (12.59; 251.19). 50% |  |
| Treatment (n. %) |  |  |  |
| No | 6 (6.1%) | 7.59 (3.98; 12.59); 37.5% | 0.009 |
| IVIg | 76 (77.6%) | 72.44 (50.12; 100.43); 39.1% |  |
| IVIg x2 | 5 (5.1%) | 32.36 (15.85; 79.43); 27% |  |
| IVIg + PLEX | 10 (10.2%) | 30.90 (12.59; 79.43); 41.8% |  |
| PLEX | 1 (1%) | 117.49 |  |

sNfL= Serum neurofilament light chain; CI=confidence interval; CV=coefficient of variation. GeoMean= geometric mean; GDS= Guillain-Barré Syndrome Disability Score; AIDP= Acute Inflammatory Demyelinating Polyneuropathy; AMAN= Acute Motor Axonal Neuropathy; AMSAN= Acute Motor-Sensory Axonal Neuropathy; MFS= Miller-Fisher Syndrome; IVIg= Intravenous Immunoglobulin; PLEX= Plasma Exchange.

**Figure 2.**
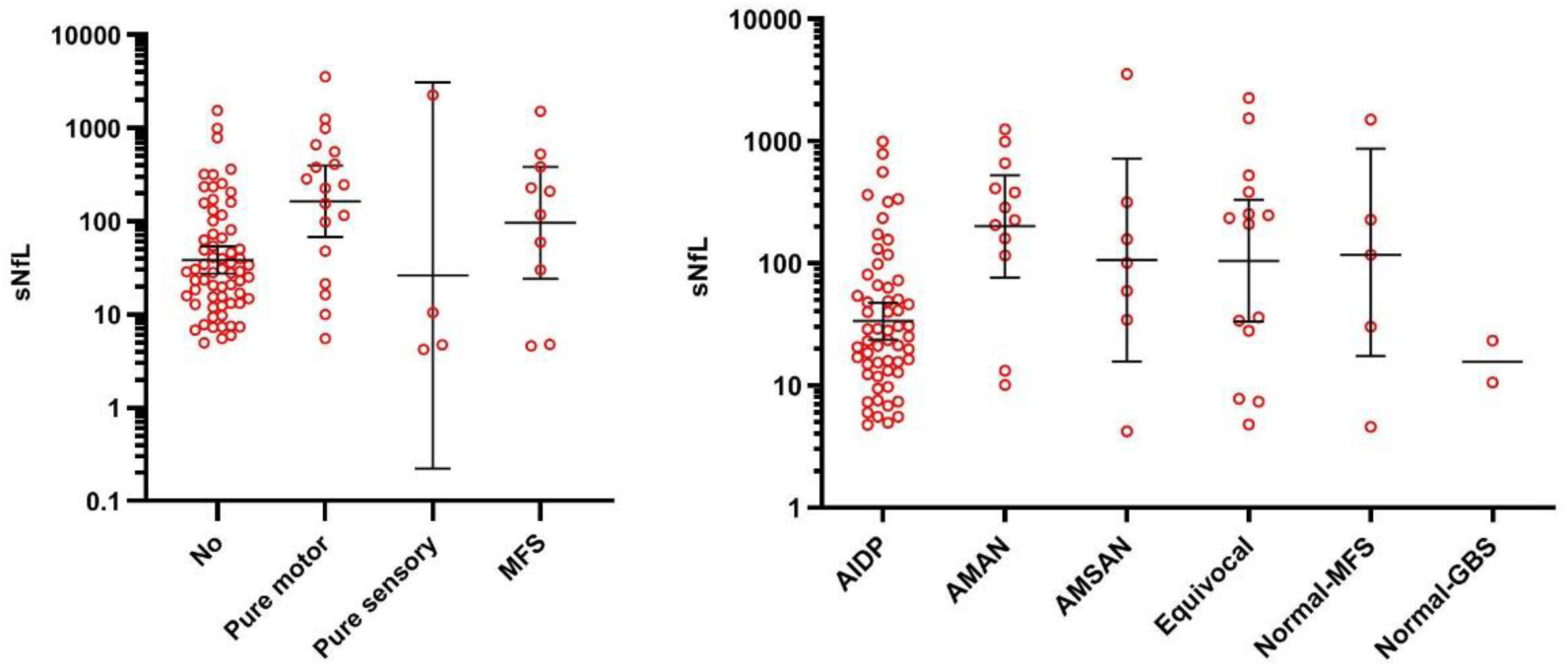
A. sNfL levels in clinical variants. B. sNfL and EMG classification. The line in the center represents the GeoMean value and the whiskers indicate the 95% confidence interval. sNfL= Serum neurofilament light chain; AIDP= Acute Inflammatory Demyelinating Polyneuropathy; AMAN= Acute Motor Axonal Neuropathy; AMSAN= Acute Motor-Sensory Axonal Neuropathy; MFS= Miller-Fisher Syndrome.

### Association of baseline sNfL with clinical scales

sNfL is clearly correlated with the GDS and R-ODS scores. sNfL levels tent to be higher with every GDS increase at inclusion time (r=0.313, p=0.002; **table 2, figure 3**). We also found a correlation with maximum GDS achieved (r=0.251, p=0.013) and GDS at all timepoints (**table 3**). Baseline sNfL levels also correlated with the R-ODS scale at 4, 26 and 52 weeks with lower R-ODS scores with higher sNfL levels (**table 3**).

**Table 3.**
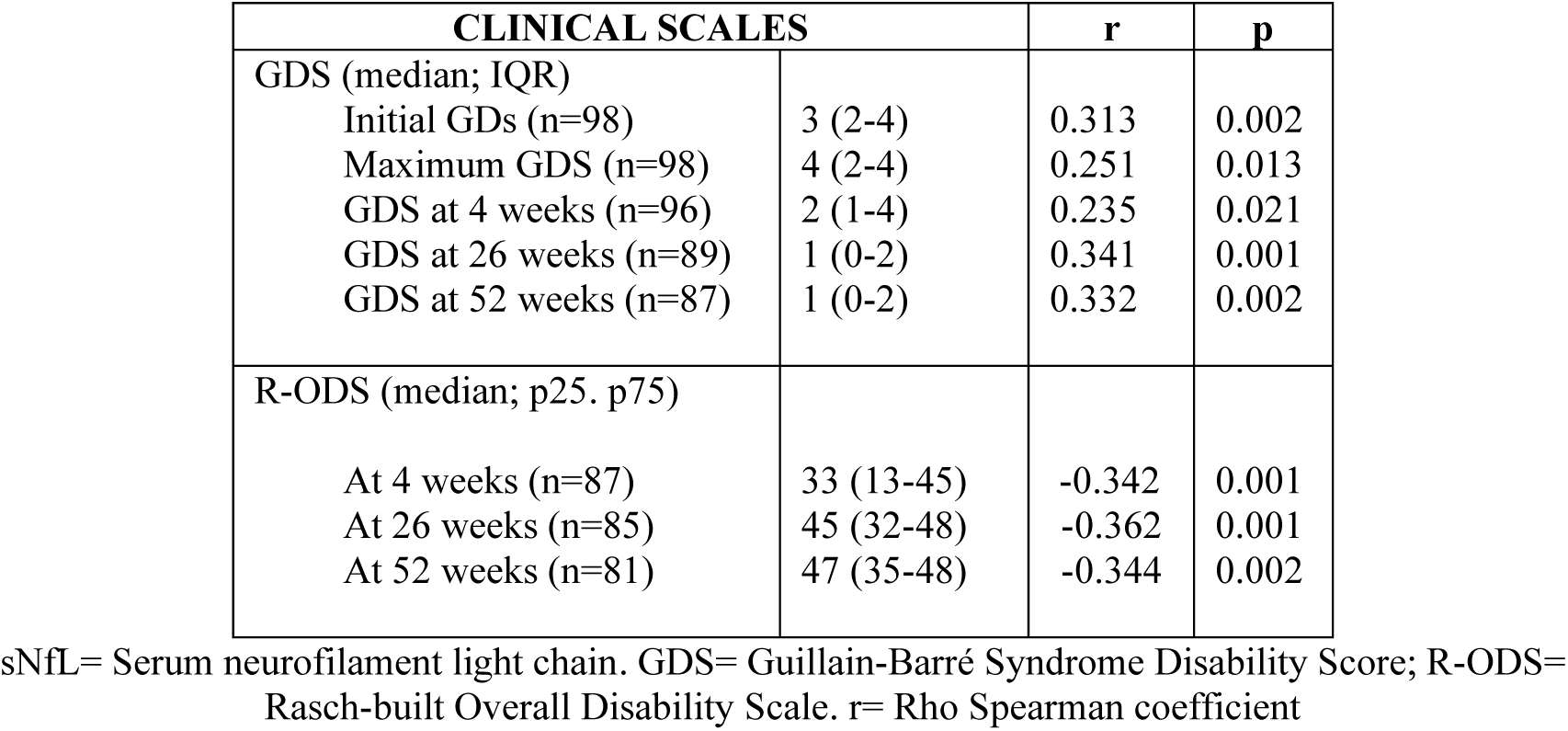
Correlation between GDS and R-ODS at different timepoints and baseline sNfL levels

| CLINICAL SCALES |  | r | p |
| --- | --- | --- | --- |
| GDS (median; IQR) |  |  |  |
| Initial GDS (n=98) | 3 (2-4) | 0.313 | 0.002 |
| Maximum GDS (n=98) | 4 (2-4) | 0.251 | 0.013 |
| GDS at 4 weeks (n=96) | 2 (1-4) | 0.235 | 0.021 |
| GDS at 26 weeks (n=89) | 1 (0-2) | 0.341 | 0.001 |
| GDS at 52 weeks (n=87) | 1 (0-2) | 0.332 | 0.002 |
| R-ODS (median; p25. p75) |  |  |  |
| At 4 weeks (n=87) | 33 (13-45) | -0.342 | 0.001 |
| At 26 weeks (n=85) | 45 (32-48) | -0.362 | 0.001 |
| At 52 weeks (n=81) | 47 (35-48) | -0.344 | 0.002 |

sNfL= Serum neurofilament light chain. GDS= Guillain-Barré Syndrome Disability Score; R-ODS= Rasch-built Overall Disability Scale. r= Rho Spearman coefficient

**Figure 3.**
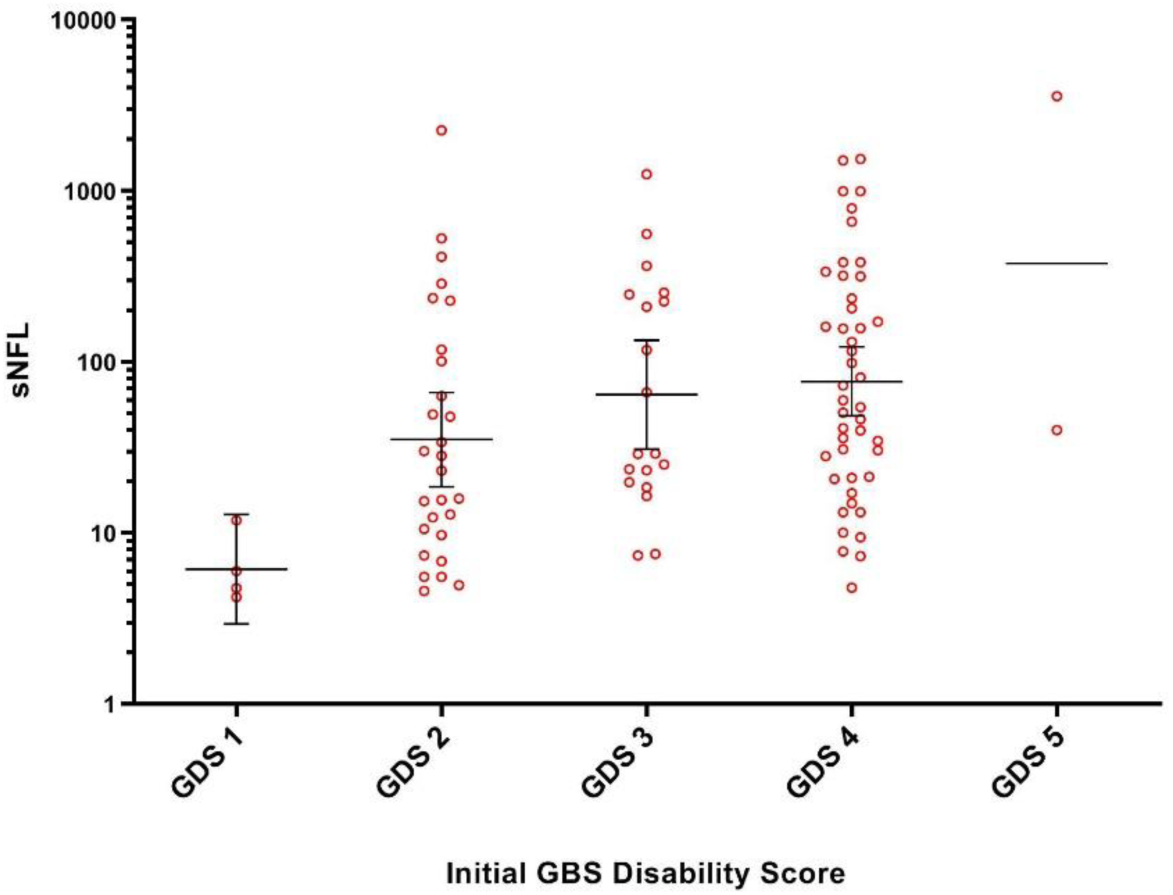
Correlation between initial GDS and baseline sNfL levels. The line in the center represents the GeoMean value and the whiskers indicate the 95% confidence interval. sNfL= Serum neurofilament light chain; GDS= Guillain-Barré Syndrome Disability Score.

### Kinetics of sNfL levels

We analyzed sNfL in 33 patients at baseline and at week 52. At one year, sNfL returned to normal control levels in all patients that had high baseline NfL levels; sNfL levels did not change in patients with normal baseline levels (**supplementary figure 2**).

### Association of baseline sNfL and prognosis

Approximately, 71% and 74.5% of patients could walk independently at 6 months and at 1 year and 60% and 67% of patients could run at 6 months and at 1 year. Ten patients needed ventilation, during a median time of 11 days (IQR 8-33), and 4 patients died. Two patients died due to respiratory insufficiency, one patient due to pneumonia and one patient because of cancer progression. Baseline sNfL levels were associated with the ability to run at 1 year (geomean sNfL levels 33.11pg/mL vs 123.03pg/mL; p<0.001). We did not find an association between baseline sNfL and ventilation (geomean sNfL levels 52.48pg/mL vs 83.17pg/mL, p=0.446) or between sNfL and death (geomean sNfL levels 53.70pg/mL vs 229.09 pg/mL, p=0.11).

In the univariate analysis, logNfL, AMAN, age and initial GDS were associated with the ability to run at 1 year and logNfL, age and initial GDS were associated with the R-ODS at 1 year (**table 4**). In the multivariable logistic regression analysis, which included 4 potential confounder variables in the initial model (age, initial GDS, AMAN and diarrhea), higher baseline NfL levels were independently associated with the inability to run at 1 year after a backward stepwise selection modelling (OR=1.65, 95% CI 1.14-2.40, p=0.009; **table 4**). Undoing the log transformation of the variable this increase represents an OR=1.019; 95% CI 1.037-1.002; for each 10pg/mL of sNfL. In the multivariable linear regression analysis to predict R-ODS, higher baseline NfL levels were also associated with less R-ODS at 1 year (β=−2.60, 95% CI −4.66-(−0.54); p=0.014) (see **table 4**).

**Table 4.** Association between baseline NfL levels and prognosis

| Univariate analysis for ability to run at 1 year |  |  |  |  |
| --- | --- | --- | --- | --- |
| Variable | OR | 95% CI | p |  |
| logNFL | 1.90 | 1.34-2.69 | <0.001 |  |
| AMAN | 6.53 | 1.57-27.17 | 0.010 |  |
| Diarrhea | 2.33 | 0.83-6.57 | 0.109 |  |
| Age | 1.04 | 1.01-1.07 | 0.023 |  |
| Initial GDS | 1.81 | 1.08-3.04 | 0.025 |  |
| Univariate analysis for R-ODS at 1 year |  |  |  |  |
| Variable | Beta coefficient | 95% CI | Std beta | p |
| logNFL | -3.67 | -5.65-(-1.69) | -0.40 | <0.001 |
| AMAN | -6.14 | -16.48-4.19 | -0.14 | 0.240 |
| Diarrhea | -6.03 | -14.41-2.34 | -0.17 | 0.155 |
| Age | -0.27 | -0.47-(-0.08) | -0.31 | 0.007 |
| Initial GDS | -5.54 | -8.63-(-2.44) | -0.39 | 0.001 |
| Multivariable logistic analysis for ability to run at 1 year* |  |  |  |  |
| Variable | OR | 95% CI | p |  |
| logNfL | 1.65 | 1.14-2.40 | 0.009 |  |
| AMAN | 6.19 | 1.01-38.02 | 0.049 |  |
| Age | 1.05 | 1.01-1.09 | 0.022 |  |
| Multivariable linear analysis for R-ODS at 1 year** |  |  |  |  |
| Variable | Beta | 95% CI | Std beta | p |
| logNfL | -2.60 | -4.66-(-0.54) | -0.29 | 0.014 |

\*Results from the multivariable logistic regression analysis and \*\*results from the multivariable linear regression analysis after a backward stepwise selection modelling. The variables included in the initial model were age, initial GDS, AMAN and diarrhea.
logNfL= log-transformed neurofilament light chain; AMAN=Acute Motor Axonal Neuropathy; GDS= Guillain-Barré Syndrome Disability Score; R-ODS= Rasch-built Overall Disability Scale; Std= Standardized. OR= Odds ratio. CI= Confidence Interval.

We additionally repeated the two multivariable analyses (logistic and linear) with and without MFS patients. We mainly focused in GBS patients only because MFS is considered a different disease and the aim of the study was to assess sNfL to predict GBS outcome. Nonetheless, multivariable analysis including MFS in the models has been also performed with very similar results (**supplementary table 1**).

### Predictive capacity of sNfL levels to predict clinically relevant outcomes

We finally evaluated the ability of baseline sNfL levels to predict the outcome in 5 clinically relevant endpoints: (1) ability to run at 1 year, (2) inability to run at 1 year, (3) inability to walk without assistance at 1 year, (4) ventilation, and (5) death. As described in Methods, we performed a ROC analysis for each endpoint selecting the cut-off points with the highest specificity. We observed that sNfL levels higher than 319pg/mL classified patients unable to walk independently at 1 year (OR=5.20, 95% CI 1.02-26.34, p=0.047, specificity 89.4%, sensitivity 33.3%); sNfL levels higher than 248pg/mL classified patients unable to run at 1 year (OR=6.81, 95% CI 1.64-28.21, p=0.008, specificity 94.2%, sensitivity 39.3%) and sNfL levels lower than 34pg/mL predicted complete recovery, defined as the ability to run at 1 year (OR=6.59, 95% CI 2.02-21.46, p=0.002, specificity 82.1%, sensitivity 69.2%). We were unable to define good prognostic sNfL level cut-off points for ventilation and death due to the low number of patients. AUC, PPV and NPV for each cut-off point are detailed in **supplementary table 2**. The 3 cut-off points values maintain their significance evaluating R-ODS at 1 year by linear regression (**supplementary table 2**).

## DISCUSSION

Our study shows that sNfL levels are a valuable biomarker in GBS patients that correlates with diverse clinical, epidemiological and electrophysiological features and that is associated with clinical outcomes (residual disability) independently of other known prognostic variables: sNfL levels are increased in patients with GBS compared to controls and correlate with disease severity and with pure motor presentation and AMAN electrophysiological variants. This study also shows that sNfL levels tested at admission can predict prognosis at 1 year and establishes different cut-off points that predict moderate disability (ability to walk independently), mild disability (inability to run) and complete recovery (ability to run) at 1 year in our sample of patients. As expected, due to the monophasic nature of GBS, all patients with high sNfL levels at baseline returned to normal levels at one year.

Baseline characteristics in GBS patients and prognostic values (ability to walk at 6 months and 1 year) in our cohort are similar to other studies and similar to the IGOS global dataset ^22^, except for an increased proportion of AMAN (12.2%) and MFS (10.2%) patients. This could be a selection bias due to the higher severity of AMAN patients or the rarity of MFS variant.

We found a correlation between baseline sNfL levels and disease severity, in agreement with previously reported findings in other peripheral neuropathies; such as chronic inflammatory demyelinating polyneuropathy (CIDP)^23^, Charcot-Marie-Tooth disease^24^ (CMT) or neuropathy in hereditary transthyretin amyloidosis^25^. The median levels of sNfL in GBS patients were higher than the sNfL levels reported in other peripheral neuropathies, but there is a high variability among GBS patients, some of them with levels 100-fold higher than HC. This variability is partially explained by the different clinical and EMG variants, being baseline sNfL levels higher in axonal variants (AMAN). Other factors, not considered in this study, could have influenced heterogeneity in sNfL levels (treatment, comorbidities, potential misdiagnosis with GBS mimics…) and will need to be assessed in larger cohorts. We also found elevated sNfL levels in patients with MFS despite their benign prognosis (87.5% were able to rum at 6 months and their median R- ODS was 48). When we analyzed the NCS for MFS patients we found five patients with normal NCS, four patients with equivocal NCS and one patient with AMSAN, classified as MFS-overlap syndrome. One possible explanation of the sNfL elevation in MFS patients despite unremarkable peripheral nerve electrophysiological alterations could be that either preganglionar nerve roots (difficult to assess with routine electrophysiology) or cerebellar damage are driving this sNfL increase instead of peripheral nerve damage^26–28^ but larger cohorts with MFS are needed.

Smaller studies have previously studied neurofilament levels^9–13^ in serum and CSF and some of them found an association of poor outcomes with severity^14^. However, we provide the first large prospective study assessing the relationship of sNfL (tested with SiMoA) with GBS features and long-term prognosis. We found a clear relationship between sNfL and prognosis: high baseline sNfL were associated with inability to run and with R-ODS scores at 1 year in a multiple logistic and linear regression models, adjusted by known prognostic factors. We also aimed to establish cut-off points to predict clinically relevant outcomes with high specificity. These three cut-off points allowed us to predict that in our sample: (1) a patient with baseline sNfL >319 pg/ml is not going to be able to walk independently at 1 year with almost 90% specificity; (2) a patient with baseline sNfL >248pg/ml is not going to be able to run at 1 year with a 94.2% of specificity; and (3) a patient with baseline sNfL< 34pg/ml is going to be able to run at 1 year with a 82.1% of specificity. Considering natural history of GBS, most patients are going to walk independently at 1 year anyway, but early recognition of patients with poor and excellent prognoses could eventually help physicians guide treatment choices in the future and provide better prognostic information for patients in the recovery phase. Incorporating sNfL in the existing prognostic models for GBS^2–4^ could be an option to improve the GBS prognosis prediction. Although our study provides one of the largest prognostic studies in GBS, larger studies should confirm our findings and establish optimized sNfL cut-off points for relevant clinical endpoints in more diverse populations and include other important confounders (treatment).

We have not found a strong correlation between the disease characteristics and NfL in CSF, as it happens in other diseases such as multiple sclerosis^29^. This could be because the immune response in GBS occurs and leads to nerve pathology predominantly in the periphery and not intrathecally. This may also explain why the sNfL and CSF NfL do not correlate with CSF protein levels. Due to the low number of patients in whom we have measured the NfL in CSF, caution should be made in interpreting our data.

An important issue in GBS and other inflammatory neuropathy trials is the lack of biomarkers that could be used as surrogate markers for disease activity and as secondary endpoints in clinical trials. Clinical trials in inflammatory neuropathies and GBS are typically performed using clinical scales as primary and secondary endpoints. Moreover, the traditional primary endpoint (ability to walk unaided at 6 months) may not be sensitive enough to detect GBS natural history modifications by assayed therapies^30^. The relationship of sNfL levels with disease severity, axonal damage and prognosis suggest that sNfL levels could be an informative secondary endpoint for phase II clinical trials or of treatment response as it happens in other diseases^31^.

One of the limitations of our study is the lack of exclusion criteria in the IGOS study. We did not collect previous neurologic diseases that could raise NfL levels in our patients. In our sample, we have seven patients that have comorbidities affecting mobility, but we did not exclude those patients from multivariable analyses to strictly follow IGOS study inclusion and exclusion criteria. Another point to study would be the time to recovery of patients with high sNfL vs patients with low sNfL. To evaluate this, it would be necessary to perform more frequent or self-registered visits informing about relevant outcomes, since we had no visits scheduled between 6 and 12 months. Furthermore, our stablished cut-off points are valid only for our sample of patients. Larger studies with repeated sampling and addressing short-term dynamics of the sNfL levels could address the relationship of sNfL levels with treatment response, best time points to test sNfL for prognostic purposes and better representative cut-off points for clinically relevant endpoints, but our study provides the proof of concept to support the use of the sNfL as a prognostic biomarker in GBS in the future.

In summary, our study proves that sNfL are increased in GBS patients, that they correlate with disease severity and axonal damage and that they could be used as an informative prognostic biomarker in GBS patients.

## Data Availability

**Abbreviations**

AIDP: Acute Inflammatory Demyelinating Polyneuropathy
AMAN: Acute Motor Axonal Neuropathy
AMSAN: Acute Motor-Sensory Axonal Neuropathy
CI: Coefficient Interval
CSF: cerebrospinal fluid
GBS: Guillain-Barré syndrome
GeoMean: geometric mean
GDS: GBS Disability Score
HC: Healthy controls
IVIg: Intravenous Immunoglobulin
logNfL: log-transformed neurofilament light chain
MFS: Miller-Fisher Syndrome
MS: Multiple sclerosis
PLEX: Plasma Exchange
sNfL: serum neurofilament light chain
(NCS): nerve conduction studies
OR: Odds ratio
R-ODS: Rasch-built Overall Disability Scale.

## Acknowledgements

We would like to thank all our patients for their patience and collaboration.

## FIGURES

**Supplementary figure 1.**
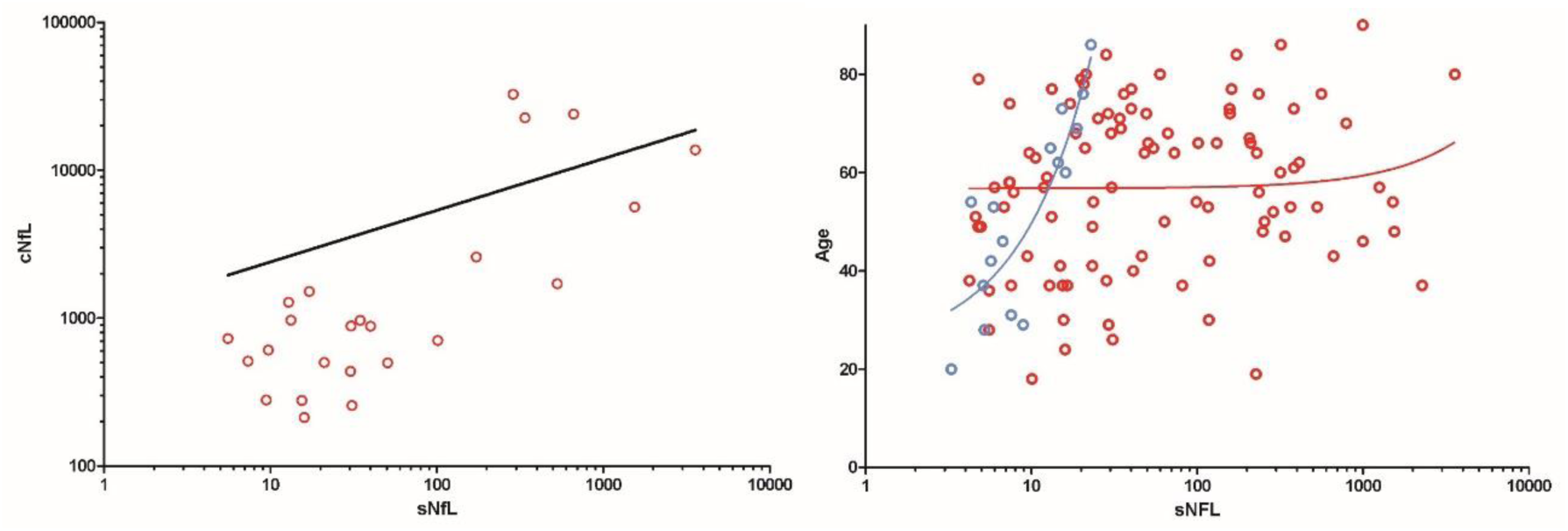
A. Correlation between sNfL and CSF. B. Correlation between sNfL and age. Dots in red represent Guillain-Barré syndrome patients, dots in blue represent healthy controls. sNfL= serum neurofilament light chain; cNfL= CSF neurofilament light chain

**Supplementary figure 2.**
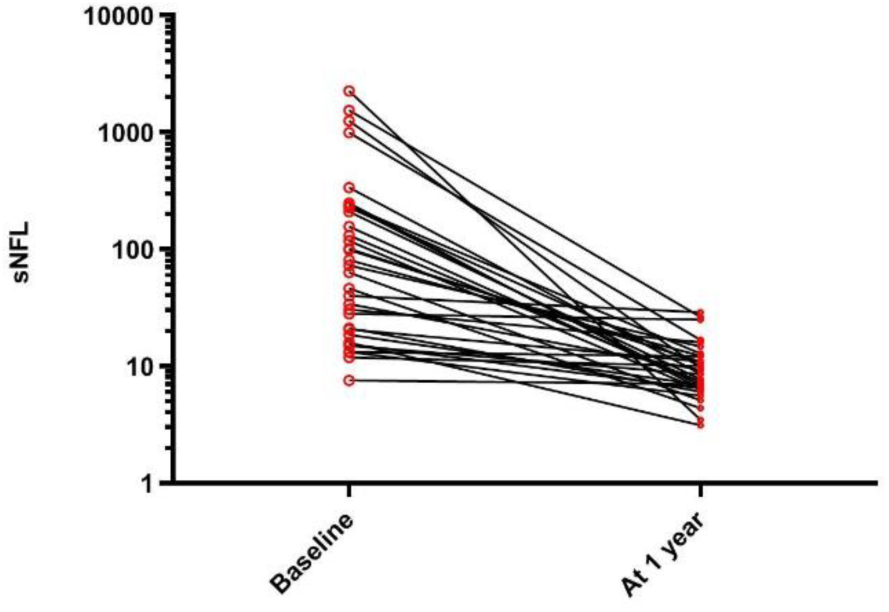
sNfL Kinetics.

**Supplementary Table 1.** Association between baseline NfL levels and prognosis including MFS patients:

| Univariate analysis for ability to run at 1 year |  |  |  |  |
| --- | --- | --- | --- | --- |
| Variable | OR | 95% CI | p |  |
| logNFL | 1.67 | 1.23-2.27 | 0.001 |  |
| AMAN | 7.23 | 1.75-29.89 | 0.006 |  |
| Diarrea | 2.34 | 0.86-6.41 | 0.097 |  |
| Age | 1.04 | 1.01-1.07 | 0.019 |  |
| Initial GDS | 1.95 | 1.17-3.25 | 0.010 |  |
| Univariate analysis for R-ODS at 1 year |  |  |  |  |
| Variable | Beta coefficient | 95% CI | Std beta | p |
| logNFL | -3.27 | -5.19-(-1.35) | -0.36 | 0.001 |
| AMAN | -6.42 | -16.17-3.93 | -0.14 | 0.220 |
| Diarrea | -5.25 | -13.42-2.92 | -0.14 | 0.205 |
| Age | -0.29 | -0.48-(-0.09) | -0.32 | 0.004 |
| Initial GDS | -5.99 | -8.93-(-3.04) | -0.42 | <0.001 |
| Multivariable logistic analysis for ability to run at 1 year* |  |  |  |  |
| Variable | OR | 95% CI | p |  |
| logNfL | 1.49 | 1.07-2.09 | 0.019 |  |
| AMAN | 8.91 | 1.49-53.14 | 0.016 |  |
| Age | 1.05 | 1.01-1.09 | 0.010 |  |
| Multivariable linear analysis for R-ODS at 1 year** |  |  |  |  |
| Variable | Beta | 95% CI | Std beta | p |
| logNfL | -2.22 | -4.12-(-0.33) | -0.24 | 0.022 |
| Initial GDS | -3.91 | -7.06- (-0.76) | -0.27 | 0.016 |

\*Results from the multivariable logistic regression analysis and \*\*results from the multivariable linear regression analysis after a backward stepwise selection modelling. The variables included in the initial model were age, initial GDS, AMAN and diarrhea.
logNfL= log-transformed neurofilament light chain; AMAN=Acute Motor Axonal Neuropathy; GDS= Guillain-Barré Syndrome Disability Score; R-ODS= Rasch-built Overall Disability Scale; Std= Standardized

**Supplementary table 2.** Clinically relevant sNfL cut-off points:

|  |  | For each clinically relevant endpoint |  |  |  |  |  |  | For evaluating R-ODS at 1 year |  |  |
| --- | --- | --- | --- | --- | --- | --- | --- | --- | --- | --- | --- |
|  |  | Multivariable Logistic Regression* |  |  | ROC Analysis |  |  |  | Multivariable Linear Regression** |  |  |
| Endpoint | Cut-off point (pg/ml) | OR (95% CI) | p | AUC | Specificity | Sensitivity | PPV | NPV | $\beta$ (95% CI) | Std $\beta$ | p |
| No ability to walk independently at 1 year | 319 | 5.20 (1.02-26.34) | 0.047 | 0.714 | 89.4% | 33.3% | 100% | 84.6% | -15.53 (-24.80-(-6.26)) | -0.37 | 0.001 |
| No ability to run at 1 year | 248 | 6.81 (1.64-28.21) | 0.008 | 0.803 | 94.2% | 39.3% | 73.3% | 73.9% | -12.25 (-21.04-(-3.46)) | -0.32 | 0.007 |
| Ability to run at 1 year | 34 | 6.59 (2.02-21.46) | 0.002 | 0.834 | 82.1% | 69.2% | 87.5% | 57.5% | 11.13 (4.75-17.50) | 0.38 | 0.001 |

\*This model includes logNfL, AMAN and Age. \*\*This model includes logNfL.
OR=Odds ratio. AUC=area under the curve; PPV=Positive Predictive Value. NPV= Negative Predictive Value; R-ODS= Rasch-built Overall Disability Scale; CI=Confidence Interval; Std=Standardized

